# Virtual and In-Person Psychiatric Care: Appointment Completion and Capacity Use in a Public Mental Health Service in Peru

**DOI:** 10.64898/2026.09.08.26362068

**Authors:** Paulo Ruiz-Grosso, Luis Macedo-Orrego, María Rivera-Encinas, María Carazas-Vera, Diego Rodríguez-Vargas, Alejandra Arosemena, Abel Sagástegui, Sonia Zevallos-Bustamante

## Abstract

**Objective:** To compare appointment completion and offered-slot utilization between virtual and in-person outpatient psychiatric care in Peru.

**Methods:** We conducted a retrospective ecological longitudinal study using routinely collected monthly aggregate administrative indicators from January 2022 through June 2026, with modality-month as the unit. Virtual-minus-in-person differences and 95% confidence intervals were estimated using Newey-West standard errors with 12 lags. June 2026 was partial; slot utilization was restricted to 2024–2026.

**Results:** Across 54 paired months, mean completion was 90.9% (SD=3.9) for virtual and 83.8% (SD=2.7) for in-person care; difference +7.2 percentage points (95% CI=4.4–9.9). Across 30 paired months, slot utilization was 73.0% (SD=8.3) and 90.5% (SD=3.8), respectively; difference −17.5 percentage points (95% CI=−24.5 to −10.4). Sensitivity analyses were similar.

**Conclusions:** Virtual care had higher completion but lower slot utilization. Attendance and capacity use should be evaluated separately when planning hybrid psychiatric services.

## Introduction

Telepsychiatry can provide clinical care comparable to face-to-face treatment in several settings (1, 2), and a recent meta-analysis found lower nonattendance with telehealth on average, although heterogeneity was substantial (3). Comparative studies of appointment attendance have reported heterogeneous findings across settings and periods. In Peru, prior studies have described implementation and acceptability of telepsychiatry among clinicians, patients, and community mental health stakeholders (4–6). The present analysis addresses a complementary operational question: how appointment completion and offered-capacity utilization evolved across virtual and in-person modalities during sustained routine hybrid psychiatric care.

The study setting was the National Institute of Mental Health Honorio Delgado–Hideyo Noguchi (INSM HD-HN), a national public psychiatric referral center in Lima, Peru. Virtual outpatient care was introduced during the pandemic and continued alongside in-person care after full on-site work resumed in April 2022. Clinicians selected modality case by case. Earlier cohort data from this setting found no evidence that telemedicine itself was associated with loss to follow-up, while other patient and service characteristics were associated with continuity of care (7). For service planning, appointment completion and use of offered capacity represent distinct operational questions: one describes attendance after scheduling, whereas the other describes how much available capacity reaches the scheduling stage.

We therefore conducted an ecological longitudinal study to characterize the operational profile of virtual relative to in-person psychiatric care from 2022 through 2026, focusing on appointment completion, offered-slot utilization, and temporal changes in the differences between modalities.

## Methods

We analyzed routinely collected aggregate monthly administrative data from MentalCom, the institutional outpatient dashboard of INSM HD-HN, from January 2022 through June 2026. June 2026 was a partial observation with an operational cutoff of June 15. The unit of analysis was modality-month. The study included adult and older-adult outpatient visits delivered by psychiatrists; no participants were recruited because the analysis was a census of eligible aggregate monthly records. Eligible records were selected using registry filters for outpatient care, the Adult and Older Adult program, psychiatrist professional category, calendar month, and modality. No patient-level diagnostic or procedural codes, free-text fields, or identification algorithms were used, and no person-level or cross-database linkage was performed. Investigators accessed only aggregate service-level registry outputs and did not access patient-level clinical records or identifiers. Virtual visits were delivered under the institution’s telework arrangement, and the registry did not distinguish telephone from video visits. Patient-level age, sex, race-ethnicity, diagnosis, illness severity, socioeconomic characteristics, and visit type were not available in the analytic data.

For each modality and month, the registry provided offered slots, scheduled appointments, and completed appointments. Appointment completion was defined as the number of completed appointments divided by the number of scheduled appointments; nonattendance was its exact complement. Slot utilization was defined as the number of scheduled appointments divided by the number of offered slots. Data cleaning excluded annual summary (“Total”) rows, verified one record per modality-month, and checked for missing values in variables required for the analyzed indicators. No duplicate modality-month records or missing values in those variables were identified. We recalculated indicators from the source counts and checked internal consistency. Completed-appointment totals reconciled exactly across modality-specific and consolidated records. Small discrepancies in scheduled-appointment totals were present in some months (maximum absolute discrepancy, 7 appointments). Offered-slot totals overlapped across modality-specific categories in 2022–2023 and therefore could not be interpreted cleanly by modality; slot utilization was analyzed only from January 2024 onward.

For each calendar month, we calculated virtual-minus-in-person differences in percentage points. We estimated mean monthly differences and 95% confidence intervals with intercept-only regressions using Newey-West standard errors with 12 lags to account for heteroskedasticity and temporal dependence over an annual cycle. We modeled temporal change in the completion difference with continuous monthly time. Sensitivity analyses used paired t tests and Wilcoxon signed-rank tests, excluded partial June 2026, used volume-weighted grouped binomial models adjusted for calendar month, and excluded January–March 2022. Analyses were run in Stata with syntax set to version 18.0 compatibility. Selected numerical results were independently cross-checked in Python 3.13.5. Reporting followed the STROBE statement and its RECORD extension for observational studies using routinely collected health data (8, 9).

The pre-project received OEAIDE approval on July 24, 2026 (OEAIDE 1054-2026) and was subsequently recorded under institutional administrative officialization code INSM-644-2026; the final administrative officialization was communicated to the research team by email on September 7, 2026. The Institutional Research Ethics Committee of INSM HD-HN determined the protocol exempt from review on August 27, 2026 (ethics registration code 644-2026; Exemption Certificate No. 015-2026-CIEI-INSM-HD-HN). Informed consent was not required because the study used only aggregate administrative indicators without patient or clinician identifiers and involved no contact with individuals. Historical records had been generated for routine operational purposes; extraction, consolidation, and research analysis occurred after the exemption determination.

## Results

After excluding 15 annual summary rows from the 177-row source extract, 162 modality-month records remained (54 each for virtual, in-person, and consolidated categories). Comparisons used 54 paired virtual and in-person months for appointment completion and 30 paired months from 2024 onward for slot utilization. Across the study period, 128,471 psychiatric visits were completed: 80,538 (62.7%) in person and 47,933 (37.3%) virtually. Because the analytic data were aggregated at service level, patient age, sex, and race-ethnicity distributions were not available.

Across 54 months, mean monthly appointment completion was 90.9% (SD=3.9) for virtual care and 83.8% (SD=2.7) for in-person care. The mean virtual-minus-in-person difference was +7.2 percentage points (95% CI=4.4–9.9), and virtual completion was higher in 51 of 54 months (Figure 1). The completion difference increased by approximately 2.2 percentage points per year (95% CI=1.5–2.8).

**Figure 1.**
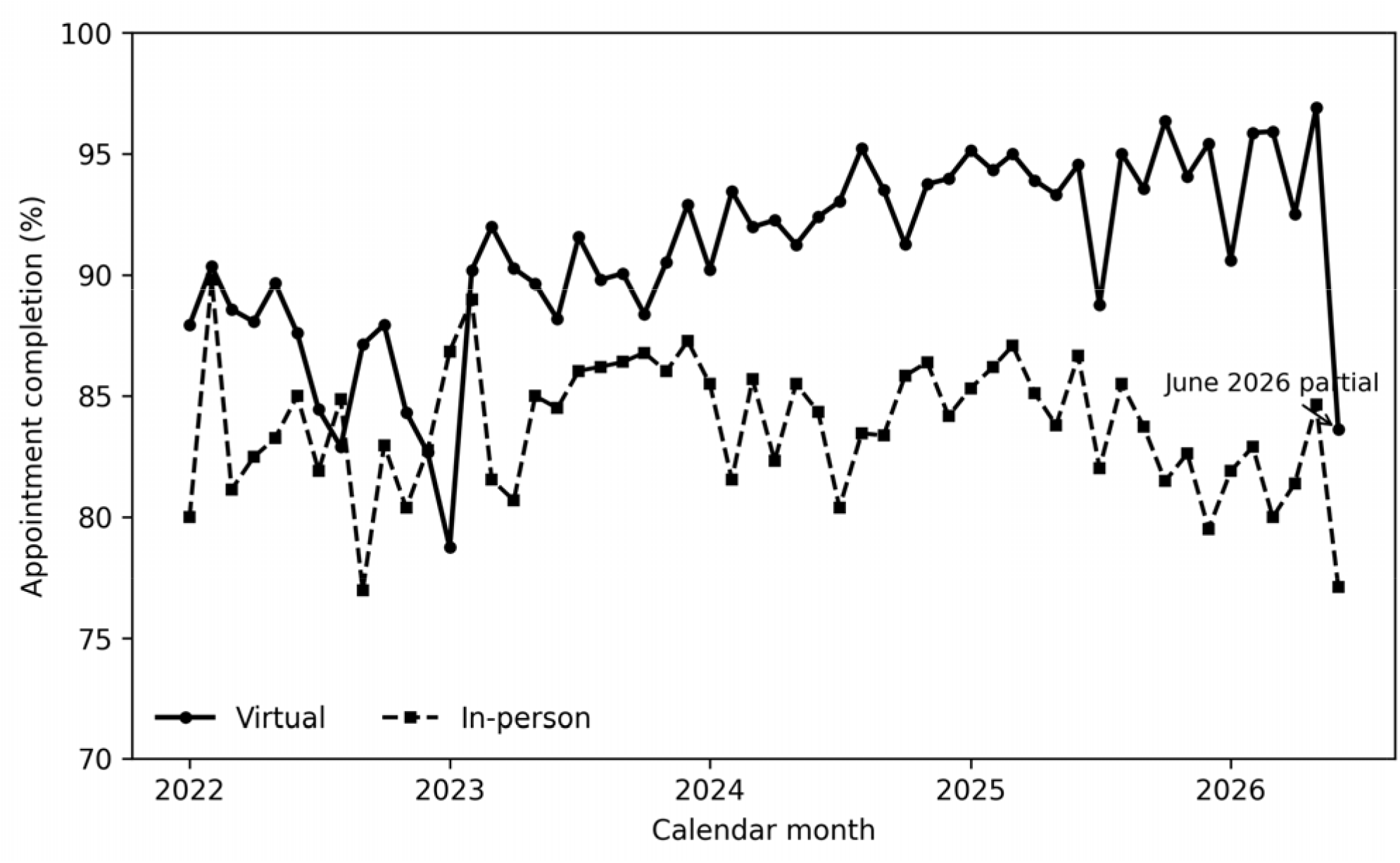
Monthly appointment completion by modality, January 2022–June 2026. The x-axis shows calendar month and the y-axis shows completed visits as a percentage of scheduled appointments. Virtual completion exceeded in-person completion in 51 of 54 paired months. June 2026 was a partial observation with an operational cutoff on June 15. The figure is descriptive; inferential estimates used Newey-West standard errors.

Across 30 months from 2024 through June 2026, mean slot utilization was 73.0% (SD=8.3) for virtual care and 90.5% (SD=3.8) for in-person care, a difference of −17.5 percentage points (95% CI=−24.5 to −10.4). Excluding partial June 2026 produced similar differences for completion (+7.2 points; 95% CI=4.4-10.0) and slot utilization (−18.1 points; 95% CI=−24.3 to −12.0). In volume-weighted analyses, model-standardized completion was 90.3% for virtual and 83.4% for in-person care (difference +6.9 percentage points; 95% CI=5.5-8.2), whereas model-standardized slot utilization was 71.8% and 90.4%, respectively (difference −18.6 percentage points; 95% CI=−21.5 to −15.8). Excluding January-March 2022 also yielded a similar completion difference (+7.3 points; 95% CI=4.4-10.1).

## Discussion

We found higher completion among scheduled virtual psychiatric appointments across more than four years of routine hybrid service delivery, whereas a smaller proportion of offered virtual slots was scheduled from 2024 onward. The attendance finding is consistent with studies reporting improved appointment adherence with telepsychiatry (10–13) and with a recent meta-analysis showing lower nonattendance with telehealth overall (3). However, heterogeneity across studies is substantial. A 2026 outpatient psychiatry study found no overall modality difference and showed that the direction of the association varied across periods (14). These results therefore describe a service-specific operational pattern rather than a general effect of virtual care.

Several limitations should be considered before interpreting these differences. Clinicians selected modality rather than assigning it randomly, creating substantial potential for confounding by indication. Patients receiving virtual care are likely to have differed in clinical stability, travel burden, preference, technology access, socioeconomic circumstances, or other factors associated with attendance. Aggregate data prevented adjustment for these characteristics and precluded individual-level inference. In addition, the registry combined telephone and video care under a single virtual-care category. Data for June 2026 were incomplete, although excluding this month did not materially change estimates. Slot utilization could be evaluated only from 2024 onward, because offered-slot counts could not be cleanly separated by modality in earlier years. Finally, findings from one national referral service may not generalize to other systems.

The study also has specific strengths. It covered 54 paired months of routine service delivery, used consistently defined administrative counts, and incorporated sensitivity analyses for temporal dependence, partial observation, and service volume. The divergence between appointment completion and slot utilization is operationally relevant. Completion describes what happened after an appointment was scheduled; slot utilization describes how much offered capacity reached the scheduling stage. Lower virtual slot utilization therefore cannot be interpreted as lower demand or inefficiency without information on scheduling rules, staffing, patient choice, and how virtual capacity was offered. Conversely, higher completion among scheduled virtual visits does not show that changing a patient from in-person to virtual care would improve attendance.

These findings add evidence from a Latin American public mental health setting, where health-system coverage and access are uneven (15). Peruvian studies have documented psychiatrist acceptability of telepsychiatry (4), patient satisfaction with telepsychiatry (5), and implementation experiences and barriers in community mental health services (6). The present study examines a complementary service-level dimension: longitudinal operational performance in routine hybrid psychiatric care. It did not measure patient experience, equity, clinical outcomes, costs, or clinician well-being. Virtual visits were commonly delivered while psychiatrists were teleworking from a designated location outside the installations of the INSM HD-HN (usually their house), so care modality cannot be separated from provider work location, an implementation dimension that may influence workflows and provider experience (16, 17). Patient-level studies are needed to characterize who accesses virtual care, determine which barriers to care are mitigated by this modality, and assess whether these administrative differences translate into improved access or clinical outcomes.

## Conclusions

In this psychiatric referral service of the public health system in Peru, scheduled virtual appointments had higher completion rates than in-person appointments, while a smaller proportion of offered virtual slots was scheduled. These noncausal aggregate findings suggest that appointment attendance and capacity utilization should be evaluated separately when planning and monitoring hybrid psychiatric services.

## Data Availability

All data produced in the present study are available upon reasonable request to the authors

## Declarations

Ethics approval and institutional administrative status: The pre-project received OEAIDE approval on July 24, 2026 (OEAIDE 1054-2026) and was subsequently recorded under institutional administrative officialization code INSM-644-2026; the final administrative officialization was communicated to the research team by email on September 7, 2026. The Institutional Research Ethics Committee of INSM HD-HN determined the protocol exempt from review on August 27, 2026 (ethics registration code 644-2026; Exemption Certificate No. 015-2026-CIEI-INSM-HD-HN). Informed consent was not required because only aggregate administrative indicators without patient or clinician identifiers were used and no individuals were contacted.

## Data availability

Requests for the study protocol, analytic code, or aggregate analytic data may be directed to the corresponding author. Access to analytic data is subject to separate institutional authorization for data release.

## Reporting guideline

Reporting followed the STROBE statement and its RECORD extension for observational studies using routinely collected health data.

## Generative AI disclosure

OpenAI ChatGPT (GPT-5.6 Sol) was used to assist with manuscript language editing, structural revision, statistical-code drafting and review, and plotting-code preparation. AI-assisted output affected the title, abstract, main-text wording, figure legend, and submission formatting. Generative AI was not used to generate, impute, or modify the source data or to make final analytical or interpretive decisions. All AI-assisted outputs were critically reviewed by the authors, and numerical results were verified against the statistical output. The authors take full responsibility for the submitted work.

